# Correlation of ECG and Cardiac MRI for Assessment of Ventricular Hypertrophy and Dilatation in Adults with Repaired Tetralogy of Fallot

**DOI:** 10.1101/2023.10.05.23296629

**Authors:** Shanjot Brar, Mehima Kang, Amit Sodhi, Marc W. Deyell, Zachary Laksman, Jason G. Andrade, Matthew T. Bennett, Andrew D. Krahn, John Yeung-Lai-Wah, Richard G. Bennett, Amanda Barlow, Jasmine Grewal, Gnalini Sathananthan, Santabhanu Chakrabarti

## Abstract

**Background:** Surgically repaired Tetralogy of Fallot (rTOF) is associated with progressive right ventricular hypertrophy (RVH) and dilation (RVD). Accurate estimation of RVH/ RVD is vital for the ongoing management of this patient population. The utility of the ECG in evaluating patients with rTOF with pre-existing right bundle branch block (RBBB) has not been studied. We aimed to determine the sensitivity/specificity of currently established ECG criteria in detecting RVH/RVD in this patient population.

**Methods:** We included consecutive patients diagnosed with rTOF who underwent CMR performed at our regional referral centre between January 2012 and December 2019. Each CMR was assessed for LVH, LVD, RVH or RVD diagnosis. The ECG corresponding to the CMR was then used to determine RVH/LVH for specificity and sensitivity analysis.

**Results:** Our study included 163 consecutive rTOF patients. The specificity for ECG-based criteria for LVH was 100.00% (95% C.I. (87.75, 100.00)), and the sensitivity was 7.19% (95% C.I. (3.15, 12.83)). When RBBB was present, specificity for RVH was 100.00% (95% C.I. (84.56, 100.00)), and sensitivity was 7.69% (95% C.I. (3.75, 13.69)). When RBBB was absent, specificity for RVH was 100.00% (95% C.I. (15.81, 100.00)), and sensitivity was 0.00% (95% C.I. (0.00, 33.63)). A regression model with the entire group of 163 ToF patients, based on the Sokolow-Lyon criterion (sum of R in V1 + S in V5/V6), produced a new suggested criterion for the diagnosis of RVH in patients with rTOF, which was a sum of R in V1 + S in V5/V6 greater than 13.25 mm. This model’s sensitivity for RVH detection was 69.1%, and specificity was 36.8%.

**Conclusions:** Standard ECG voltage criteria have poor sensitivity for detecting right and left ventricular chamber hypertrophy and dilatation in patients with rTOF, so current ECG criteria should not be used to monitor RVH/RVD in this patient population.

## INTRODUCTION

Patients with surgically repaired tetralogy of Fallot (rTOF) often experience progressive right ventricular (RV) remodelling and dilation.^1^ Longitudinal studies show that RV remodelling stabilizes in adolescence and early adulthood, but some patients may experience rapid progression of RV dilation in later adulthood, leading to adverse outcomes.^2^ Monitoring RV size and function using cardiac MRI (CMR) is recommended from adolescence, but this is not always feasible due to resources and patient anxiety.^3^ CMR is also not feasible in patients with non-MRI conditional devices such as pacemakers and defibrillators. Furthermore, assessing right ventricular hypertrophy/dilation (RVH/RVD) through conventional imaging with echocardiography is challenging due to the complex RV shape and difficulty in measuring RV-free wall motion.^4^

Assessing cardiac chamber enlargement and hypertrophy using routine diagnostics like electrocardiogram (ECG) could benefit clinicians and patients. ECGs are widely available, inexpensive and provide valuable information on structural heart disease. In this study, we aim to determine the diagnostic accuracy of established ECG criteria in detecting both RVH/RVD or left ventricular hypertrophy/dilation (LVH/LVD) on CMR in the adult congenital heart disease (ACHD) population, specifically for patients with rTOF who often have right bundle branch block (RBBB).

## METHODS

### Study overview

We conducted a retrospective cohort study of patients with rTOF from our ACHD clinic (Pacific Adult Congenital Heart Centre, St. Paul’s Hospital, Vancouver, British Columbia, Canada) who underwent a cMRI scan from January 2012 to December 2019. Our clinic is a provincial referral center for patients with confirmed or suspected adult congenital heart disease diagnoses, drawn from a population of 5.1 million residents. This study was approved by the University of British Columbia (UBC) research ethics board, REB H21-00615.

### Cardiac MRI data

A secure, view-only electronic health record (CareConnect) was used to access patient CMR reports. All patients underwent multiplanar, multisequence, unenhanced and gadolinium-enhanced CMR on a 1.5-T platform (HDxT, GE Healthcare) according to a standard Tetralogy of Fallot protocol. LVH was determined using both left ventricular wall thickness and LV mass index. Similarly, RVH was determined using right ventricular wall thickness and RV mass index. Left/right ventricular dilation was determined using LV/RV end-diastolic diameter and LV/RV volume index. Published reference ranges for CMR in adults were utilized.^5^ For patients with repeat CMRs, we included the first available CMR on record.

### ECG data

The closest preceding ECG to each corresponding CMR was then used to determine RVH/LVH for sensitivity and specificity analysis. We used the Sokolow-Lyon and Cornell voltage criteria for the diagnosis of LVH and the Sokolow-Lyon and Myers’ voltage criteria for diagnosis of RVH.^6–8^ If the ECG met either criteria, a diagnosis of LVH or RVH was made. A diagnosis of RBBB or left bundle branch block (LBBB) was made using the criteria provided in the supplemental material (S1). Automated records were used to determine the cardiac intervals and QRS width.

### Definitions

CMRs meeting criteria for RVH/RVD diagnosis were used to establish underlying RVH/RVD for sensitivity and specificity analysis. Patients with RVH/RVD on CMR and ECG diagnosis of RVH/RVD were called true positives. Patients without RVH/RVD on CMR but no ECG diagnosis of RVH/RVD were called true negatives. Patients with RVH/RVD on CMR but no ECG diagnosis of RVH/RVD were called false negatives. Patients with no RVH/RVD on CMR but ECG diagnosis of RVH/RVD were called false positives. A similar process was repeated for patients with LVH/LVD on CMR to establish a diagnosis of underlying LVH/LVD for sensitivity and specificity analysis.

As there are no established criteria for the presence of RVH in patients with RBBB, we also performed a regression model with the entire group of 163 rTOF patients based on the Sokolow-Lyon criterion (R in V1 + S in V5/V6) and Meyer’s criterion (R/S in V1, R/S in V5/V6, and R in V1).

### Statistical analysis

The statistical analyses were completed using the IBM SPSS statistics software, version 27 (IBM Corp, Armonk, NY). Pearson chi-squared tests were used to compare categorical variables, and independent sample t-tests were used to compare continuous variables. Sensitivity and specificity analyses were completed on MedCalc statistical software.^9^ Sensitivity, specificity, disease prevalence, positive and negative predictive value, and accuracy are expressed as percentages.

Confidence intervals for sensitivity, specificity and accuracy are “exact” Clopper-Pearson confidence intervals. Confidence intervals for the predictive values are the standard logit confidence intervals given by Mercaldo et al.; except when the predictive value is 0 or 100%, a Clopper-Pearson confidence interval is reported.^10^

## RESULTS

### Baseline characteristics

Seven hundred seventy-one consecutive patients were identified as having both a diagnosis of congenital heart disease and a CMR between Jan 2012 – Dec 2019. Of these patients, a total of 163 patients had a diagnosis of rTOF and were included in our study. The mean age was 40.1 ± 11.5 years. Female patients accounted for 40.5% of the study cohort. The mean RV EF was 40.2% ± 8.7, and the mean RV volume index was 128.0 ml/m2 ± 43.9. The mean LV EF was 56.2% ± 8.3, and the mean LV volume index was 80.6 ml/m2 ± 23.8. The mean QRS duration was 131.1ms ± 33.1 (Table 1).

**Table 1.** Tetralogy of Fallot cohort baseline statistics.

|  |  |
| --- | --- |
|  | **Tetralogy of Fallot** |
|  | N=163 |
| **Baseline characteristics<sup>1</sup>** |  |
| Age<sup>2</sup> | 40.1 (11.5) |
| Male | 97 (59.5) |
| RVEF (%)<sup>2</sup> | 40.2 (8.7) |
| RV Volume Index (ml/m<sup>2</sup>)<sup>2</sup> | 128.0 (43.9) |
| LVEF (%)<sup>2</sup> | 56.2 (8.3) |
| LV Volume Index (ml/m<sup>2</sup>)<sup>2</sup> | 80.6 (23.8) |
| QRS duration (ms)<sup>2</sup> | 131.1 (33.1) |
| **TOF repair subtype<sup>1</sup>** |  |
| Unknown | 61 (37.4) |
| External conduit | 5 (3.1) |
| Outflow tract patch | 41 (25.2) |
| Transannular patch | 53 (32.5) |
| Without patch | 3 (1.8) |
| <sup>1</sup>Data presented as: n (%) unless otherwise specified; <sup>2</sup>data presented as: mean (SD)<br>TOF; tetralogy of Fallot RVEF; right ventricular ejection fraction, RV; right ventricle, LVEF; left ventricular ejection fraction, LV; left ventricle. |  |

### MRI/ECG characteristics

One hundred thirty-nine patients had RVH/RVD confirmed on CMR, seven patients had CMR proven LVH/LVD. Fourteen patients had an ECG diagnosis of RVH, and five had LVH. One hundred fifty-two patients had a diagnosis of RBBB, and zero had a diagnosis of LBBB (Table 2).

**Table 2.**
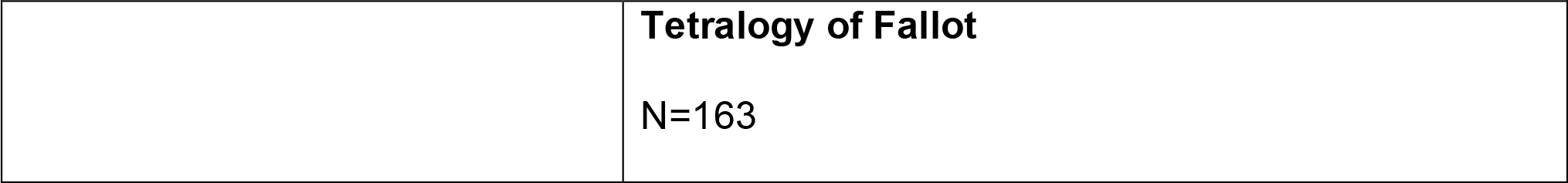
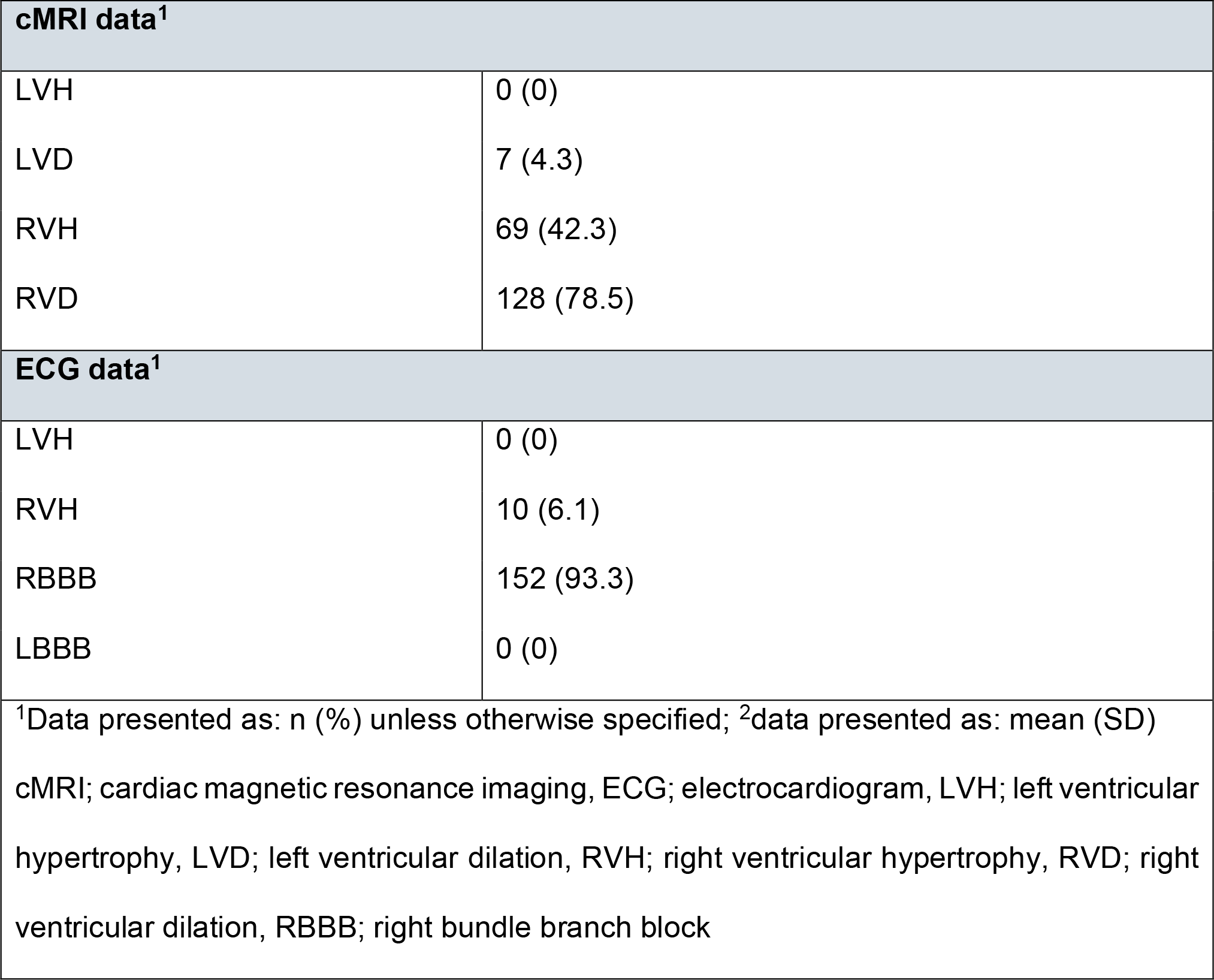
cMRI and ECG diagnosis of Tetralogy of Fallot cohort.

|  |  |
| --- | --- |
|  | **Tetralogy of Fallot** |
|  | N=163 |

| cMRI data<sup>1</sup> |  |
| --- | --- |
| LVH | 0 (0) |
| LVD | 7 (4.3) |
| RVH | 69 (42.3) |
| RVD | 128 (78.5) |
| ECG data<sup>1</sup> |  |
| LVH | 0 (0) |
| RVH | 10 (6.1) |
| RBBB | 152 (93.3) |
| LBBB | 0 (0) |
| <sup>1</sup>Data presented as: n (%) unless otherwise specified; <sup>2</sup>data presented as: mean (SD)<br>cMRI; cardiac magnetic resonance imaging, ECG; electrocardiogram, LVH; left ventricular hypertrophy, LVD; left ventricular dilation, RVH; right ventricular hypertrophy, RVD; right ventricular dilation, RBBB; right bundle branch block |  |

### Sensitivity/specificity analysis

For ECG-reported diagnosis, the specificity for LVH/LVD was 100.00% (95% CI (87.75, 100.00)), and the sensitivity was 7.19% (95% CI (3.50, 12.83)). When RBBB was absent, ECG sensitivity was 0.00% (95% CI (0.00, 33.63)). ECG specificity was 100.00% (95% CI (15.81, 100.00)). In subjects with RBBB, specificity for RVH was 100.00% (95% CI (84.56, 100.00)), and sensitivity was 7.69% (95% CI (3.75, 13.69)). (Table 3).

**Table 3.** Sensitivity, specificity, PPV and NPV of the ECG in detecting right and left ventricular chamber hypertrophy and dilatation on cardiac MRI in adults with repaired tetralogy of Fallot.

|  | Specificity (%) | Sensitivity (%) |
| --- | --- | --- |
| LVH/LVD | 100.00<br>[87.75-100.00] | 7.19<br>[3.50-12.83] |
| RVH/RVD,<br>without RBBB | 100.00<br>[15.81-100.00] | 0.00<br>[0.00-33.63] |
| RVH/RVD,<br>with RBBB | 100.00<br>[84.56-100.00] | 7.69<br>[3.75-13.69] |
| PPV; positive predictive value, NPV; negative predictive value, ECG;<br>electrocardiogram, LVH; left ventricular hypertrophy, LVD; left ventricular dilation,<br>RVH; right ventricular hypertrophy, RVD; right ventricular dilation, RBBB; right<br>bundle branch block |  |  |

The regression model with the entire group of 163 ToF patients, based on the Sokolow-Lyon criterion (R in V1 + S in V5/V6), produced new suggested criteria for the diagnosis of RVH in patients with RBBB, which was a Sokolow-Lyon sum greater than 13.2 mm. This model’s sensitivity for RVH detection was 69.1%, and the specificity was 36.8%. The regression model with the entire group of 163 ToF patients, based on the Meyers et. al criteria (R/S in V1, R/S in V5/V6, and R in V1) resulted in a negative correlation for the prediction of RVH and was therefore not applied.

## DISCUSSION

We anticipate a significant increase in our ACHD follow-up programs, particularly for patients with rTOF, as their outcomes continue to improve.^11^ While their survival rate in early childhood is excellent, morbidity remains high as they reach middle age.^12^ Risk assessment in rTOF continues to be a topic of investigation, and imaging markers of adverse right ventricular remodeling include RV dilation and increasing RV mass.^3^ These imaging markers, however are currently only available using CMR.^4^ Although there are published guidelines for routine imaging follow up of patients with rTOF, some patients do require expediated imaging referral and assessment by local experts.^4^ Monitoring disease progression is imperative in patients with rTOF as a referral for pulmonary valve replacement (PVR) is recommended before the deterioration in RVEF occurs, as RV dysfunction has shown to correlate poorly with postoperative outcomes.^13^ However, due to the growing number of follow-ups in the current environment, many patients now receive shared care with primary care physicians, internists, and community cardiologists. To aid community cardiologists in monitoring rTOF patients, establishing ECG criteria to detect chamber hypertrophy/enlargement would be helpful and could allow for more urgent referrals for CMR if necessary.

In 2009, the AHA/ACCF/HRS published standardized criteria for interpreting ECGs, including RVH. There is however, no published criteria for the detection of RVH in the presence of RBBB.^14^ This criterion was derived from studies that used cadaveric dissection of patients. These studies were small, mainly in patients with previously diagnosed RV pathology and not in the presence of RBBB.^7,15–17^ Validation of this criteria using CMR as the gold standard in the general population has shown that most ECG criteria for RVH have low sensitives (<10%) and, therefore should not be used as a screening tool.^18^ As these validation studies were done in patients without cardiovascular disease, the findings do not apply to a population at increased risk for RVH, and the AHA/ACCF/HRS concludes that the greatest accuracy of ECG RVH criteria is in patients with congenital heart disease.^14^

In this study, we analyzed the sensitivity and specificity of established ECG criteria in detecting chamber enlargement or hypertrophy, with correlation from CMR, in adults with rTOF. While we initially included all patients with congenital heart disease, we focused our analysis on a subgroup of patients with rTOF due to their anatomical homogeneity. The majority of patients in our study had RBBB, as is expected for rTOF. Our findings indicate that currently established ECG criteria have high specificity for detecting right and left ventricular chamber enlargement or hypertrophy in rTOF patients. However, they have poor sensitivity for detecting LVH/LVD and even worse sensitivity for detecting RVH/RVD. The presence of a right bundle branch block does not affect the sensitivity of the ECG in detecting RVH/RVD.

It is important to note that different phenotypes for patients with rTOF exist depending on their surgical history. Transannular patch repair typically results in RV dilatation with eccentric hypertrophy and RV-PA conduit repair typically results in lower volumes and concentric hypertrophy.^3^ Our study did not differentiate between types of repair, and it is feasible that this would result in a more accurate analysis.

These findings suggest that similar to screening in a healthy population, the surface ECG alone is insufficient for excluding progressive RVH/RVD in rTOF patients CMR should be used for routine follow-up and symptomatic patients. However, since CMR is resource-intensive, new ECG criteria for diagnosing RVD/RVH in rTOF patients are needed. We propose utilizing a sum of R wave amplitude in lead V1 and S wave amplitude in leads V5/V6 greater than 13.2 mm as a reliable indicator of RVH in this patient population, however this requires external validation.

## CONCLUSION

The current ECG voltage criteria are insufficient in detecting right ventricular hypertrophy and dilatation in rTOF patients. Despite the limitations of our study due to phenotypic variation in congenital heart disease, our findings strongly indicate the necessity for new ECG criteria to be implemented for follow-up of rTOF patients. We propose a new ECG standard: R in V1 + S in V5/V6 sum > 13.2 mm to detect RVH in the rToF.

## Data Availability

The authors confirm that the data supporting the findings of this study are available within the article and its supplementary materials.

## ABBREVIATIONS

Non-standard Abbreviations and Acronyms

rTOF: repaired tetralogy of Fallot
RV: right ventricular
CMR: cardiac MRI
ECG: electrocardiogram
RVH: right ventricular hypertrophy
RVD: right ventricular dilation
RBBB: right bundle branch block
ACHD: adult congenital heart disease
LVH: left ventricular hypertrophy
LVD: left ventricular dilation
LBBB: left bundle branch block
PVR: pulmonary valve replacement

---

**Figure 1.**
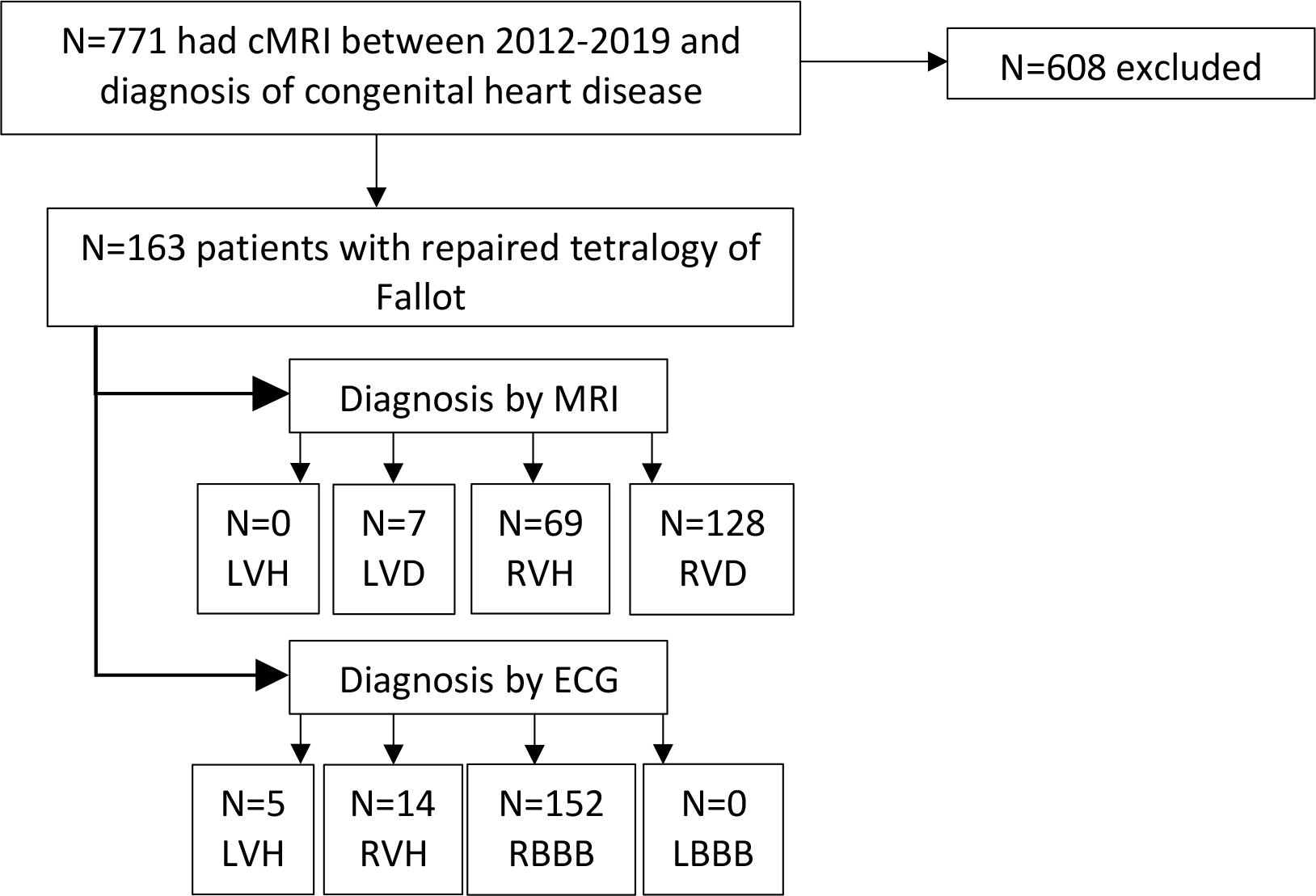

## Supplemental Material

LBBB Criteria:

- QRS duration > 120ms.
- Dominant S wave in V1, no R wave in lead V1.
- Broad monophasic R wave in lateral leads (I, aVL, V5-6).
- Absence of Q waves in lateral leads.
- Prolonged R wave peak time > 60ms in leads V5-6.

RBBB Criteria:

- QRS duration > 120ms
- RSR’ pattern in V1, with second R wave bigger than the first
- Wide, slurred S wave in lateral leads (I, aVL, V5-6)

## REFERENCES

1. Rachel M Wald, Anne Marie Valente, Kimberlee Gauvreau, Sonya V Babu-Narayan, Gabriele Egidy Assenza, Jenna Schreier, Michael A Gatzoulis, Philip J Kilner, Zeliha Koyak, Barbara Mulder, Andrew J Powell, Tal Geva. Cardiac magnetic resonance markers of progressive RV dilation and dysfunction after tetralogy of Fallot repair. Heart. 2015;101(21):1724.

2. Mertens LL. Right ventricular remodelling after tetralogy of Fallot repair: new insights from longitudinal follow-up data. European Heart Journal - Cardiovascular Imaging. 2017;18(3):371–372.

3. Valente AM, Geva T. How to Image Repaired Tetralogy of Fallot. Circulation: Cardiovascular Imaging. 2017;10(5):e004270.

4. Valente AM, Cook S, Festa P, Ko HH, Krishnamurthy R, Taylor AM, Warnes CA, Kreutzer J, Geva T. Multimodality Imaging Guidelines for Patients with Repaired Tetralogy of Fallot: A Report from the American Society of Echocardiography. Journal of the American Society of Echocardiography. 2014;27(2):111–141.

5. Kawel-Boehm N, Hetzel SJ, Ambale-Venkatesh B, Captur G, Francois CJ, Jerosch-Herold M, Salerno M, Teague SD, Valsangiacomo-Buechel E, van der Geest RJ, Bluemke DA. Reference ranges (“normal values”) for cardiovascular magnetic resonance (CMR) in adults and children: 2020 update. Journal of Cardiovascular Magnetic Resonance. 2020;22(1):87.

6. Devereux RB, Casale PN, Eisenberg RR, Miller DH, Kligfield P. Electrocardiographic detection of left ventricular hypertrophy using echocardiographic determination of left ventricular mass as the reference standard. Journal of the American College of Cardiology. 1984;3(1):82–87.

7. Myers GB, Klein HA, Stofer BE. The electrocardiographic diagnosis of right ventricular hypertrophy. American Heart Journal. 1948;35(1):1–40.

8. Sokolow M, Lyon TP. The ventricular complex in left ventricular hypertrophy as obtained by unipolar precordial and limb leads. American Heart Journal. 1949;37(2):161–186.

9. Anon. MedCalc Software Ltd. Diagnostic test evaluation calculator.

10. Mercaldo ND, Lau KF, Zhou XH. Confidence intervals for predictive values with an emphasis to case–control studies. Statistics in Medicine. 2007;26(10):2170–2183.

11. Bhat M, Mercer-Rosa L, Fogel MA, Harris MA, Paridon SM, McBride MG, Shults J, Zhang X, Goldmuntz E. Longitudinal changes in adolescents with TOF: implications for care. European Heart Journal - Cardiovascular Imaging. 2017;18(3):356–363.

12. Mark Dennis, Ben Moore, Irina Kotchetkova, Lynne Pressley, Rachael Cordina, David S Celermajer. Adults with repaired tetralogy: low mortality but high morbidity up to middle age. Open Heart. 2017;4(1):e000564.

13. Geva T, Gauvreau K, Powell AJ, Cecchin F, Rhodes J, Geva J, Del Nido P. Randomized Trial of Pulmonary Valve Replacement With and Without Right Ventricular Remodeling Surgery. Circulation. 2010;122(11_suppl_1).

14. Hancock EW, Deal BJ, Mirvis DM, Okin P, Kligfield P, Gettes LS. AHA/ACCF/HRS Recommendations for the Standardization and Interpretation of the Electrocardiogram: Part V: Electrocardiogram Changes Associated With Cardiac Chamber Hypertrophy: A Scientific Statement From the American Heart Association Electrocardiography and Arrhythmias Committee, Council on Clinical Cardiology; the American College of Cardiology Foundation; and the Heart Rhythm Society: Endorsed by the International Society for Computerized Electrocardiology. Circulation. 2009;119(10).

15. Murphy ML, Thenabadu PN, De Soyza N, Doherty JE, Meade J, Baker BJ, Whittle JL. Reevaluation of electrocardiographic criteria for left, right and combined cardiac ventricular hypertrophy. The American Journal of Cardiology. 1984;53(8):1140–1147.

16. Murphy ML, Thenabadu PN, Blue LR, Meade J, De Soyza N, Doherty JE, Baker BJ. Descriptive characteristics of the electrocardiogram from autopsied men free of cardiopulmonary disease—A basis for evaluating criteria for ventricular hypertrophy. The American Journal of Cardiology. 1983;52(10):1275–1280.

17. Butler PM, Leggett SI, Howe CM, Freye CJ, Hindman NB, Wagner GS. Identification of electrocardiographic criteria for diagnosis of right ventricular hypertrophy due to mitral stenosis. The American Journal of Cardiology. 1986;57(8):639–643.

18. Whitman IR, Patel VV, Soliman EZ, Bluemke DA, Praestgaard A, Jain A, Herrington D, Lima JAC, Kawut SM. Validity of the Surface Electrocardiogram Criteria for Right Ventricular Hypertrophy. Journal of the American College of Cardiology. 2014;63(7):672–681.

